# Evaluating the Sensitivity of SARS-CoV-2 Infection Rates on College Campuses to Wastewater Surveillance

**DOI:** 10.1101/2020.10.09.20210245

**Authors:** Tony E. Wong, George M. Thurston, Nathaniel Barlow, Nathan Cahill, Lucia Carichino, Kara Maki, David Ross, Jennifer Schneider

## Abstract

As college campuses reopen, we are in the midst of a large-scale experiment on the efficacy of various strategies to contain the SARS-CoV-2 virus. Traditional individual surveillance testing via nasal swabs and/or saliva is among the measures that colleges are pursuing to reduce the spread of the virus on campus. Additionally, some colleges are testing wastewater on their campuses for signs of infection, which can provide an early warning signal for campuses to locate COVID-positive individuals. However, a representation of wastewater surveillance has not yet been incorporated into epidemiological models for college campuses, nor has the efficacy of wastewater screening been evaluated relative to traditional individual surveillance testing, within the structure of these models. Here, we implement a new model component for wastewater surveillance within an established epidemiological model for college campuses. We use a hypothetical residential university to evaluate the efficacy of wastewater surveillance to maintain low infection rates. We find that wastewater sampling with a 1-day lag to initiate individual screening tests, plus completing the subsequent tests within a 4-day period can keep overall infections within 5% of the infection rates seen with traditional individual surveillance testing. Our results also indicate that wastewater surveillance can be an effective way to dramatically reduce the number of false positive cases by identifying subpopulations for surveillance testing where infectious individuals are more likely to be found. Through a Monte Carlo risk analysis, we find that surveillance testing that relies solely on wastewater sampling can be fragile against scenarios with high viral reproductive numbers and high rates of infection of campus community members by outside sources. These results point to the practical importance of additional surveillance measures to limit the spread of the virus on campus and the necessity of a proactive response to the initial signs of outbreak.

**Author Summary:** College campuses have employed a variety of measures to keep their communities safe amid the SARS-CoV-2 pandemic. Many colleges are implementing surveillance testing programs wherein students are randomly selected to be tested for SARS-CoV-2. These strategies aim to manage the number of infections among the student population by isolating infected individuals. Some colleges are monitoring wastewater on their campuses for signs of the virus, which has been found to be capable of detecting viral RNA. If a wastewater sample shows signs of viral RNA, then screening tests are administered to the individuals who live or work in the buildings that contribute to the sewer in question. We present a model for such wastewater surveillance within a larger model for the spread of SARS-CoV-2 on a college campus. We show that wastewater surveillance can reduce the number of false positive cases and the associated disruptions to student life, while maintaining similar overall numbers of infections. However, we find that surveillance testing strategies that rely solely on wastewater sampling may be less effective if the local transmission rate of the virus is high, or if the rate of infection of members of the campus community by outside sources is high.

## 1 Introduction

As colleges put into action their reopening plans for Fall 2020, a natural experiment in epidemic management is unfolding. This experiment is stress-testing colleges’ strategies for reducing the spread of SARS-CoV-2 on their campuses. These strategies include reducing the capacities of classrooms and residence halls, mandating the use of face masks, implementing extensive sanitization and cleaning protocols between classes, putting in place social distancing requirements, and banning large gatherings. Additionally, many colleges are using traditional individual screenings to test students for the virus [1–3] at periodic intervals or by random sampling throughout the fall term. Previous work has found that this type of surveillance testing is critical for controlling the spread of virus on campus [4–6], and that surveillance testing the campus population every 2-10 days is a minimum requirement to keep the overall number of infections manageable.

Such frequent screenings of all of a university’s students places a high financial and logistical burden on universities, as well as an intrusion on the lives and comfort of students. As an alternative or supplementary form of viral surveillance, many municipalities and universities are turning to collecting and testing wastewater from their campuses for signs of viral ribonucleic acid (RNA) [7]. Viral presence in wastewater can be a leading indicator for positive cases in traditional individual screening tests [8–10]. In this approach, wastewater is collected at sewer locations around campus and tested in a laboratory, typically by a testing method that incorporates a polymerase-chain reaction (PCR) assay [11]. If the wastewater sample from a particular sewer shows signs of viral presence, then the individuals in the building(s) that the wastewater sampling location serves can be given traditional individual screening tests for SARS-CoV-2. In this way, wastewater surveillance can be an effective tool to reduce the overall number of randomized screening tests required in order to maintain low infection rates. However, the epidemiological models used to inform college campus reopening strategies do not yet include a representation of wastewater surveillance. In this work, we present a wastewater module for the susceptible-exposed-infected-recovered (SEIR) model of Paltiel et al. [4].

The SEIR modeling approach [12] employs a type of compartment model, wherein all individuals are assumed to be in one of four general categories: susceptible to infection, exposed (but not yet infectious or symptomatic), infected (either symptomatic or asymptomatic), and removed/recovered (recovered from the infection or deceased) [13]. These models do not track individual persons (i.e., are not agent-based), and as such SEIR models are simple and stylized in their representation of persons and processes. However, this simplicity leads to high computational efficiency, making SEIR models ideal for the large numbers of model simulations required for uncertainty and sensitivity analyses, and for informing decision-making to manage campus community health under uncertain conditions. Such decision support is critical when campus officials must manipulate decision levers including the rate at which to administer screening tests to the campus population and what share of courses should be held on-campus versus online. Previous efforts to account for the sensitivity of projected infection rates have focused on simple Monte Carlo sensitivity analyses, wherein each parameter is sampled from a probability distribution and the effects of these changes on the infection rates is compared (e.g., 5). These foundational approaches assess the sensitivity of infection counts to uncertainties in model parameters, which represent real, on-the-ground uncertainties. However, previous work provides a largely qualitative view of these sensitivities; a formal global sensitivity analysis is still needed. Such a global analysis would quantify how the variation in infection rates is attributable to each uncertain model parameter and potential decision lever, including, for example, the viral reproduction rate and the rate at which screening tests are administered to the campus population.

Here, we address these issues by assigning marginal prior probability distributions to each uncertain model input parameter. We sample from these prior distributions to examine the sensitivity of infection rate to uncertainty in the model parameters. By considering changes in these parameters in combination, we conduct a formal global sensitivity analysis to attribute the variance in the total number of infections to variation in each of the input parameters (including decision levers) and interactions among the parameters. We use a previously published SEIR model to incorporate a model component to represent wastewater surveillance testing. We use our new model to assess the ability of a wastewater surveillance system to prevent numbers of infections from exceeding a desired maximum. We examine the vulnerability of a wastewater surveillance system to failures in underlying assumptions, including increases in the viral reproductive rate, larger numbers of new infections of campus members from outside sources, and higher rates of noncompliance with quarantine procedures.

## 2 Methods

### 2.1 SEIR model

Our model is a modified version of the SEIR model of Paltiel et al. [4]. The interested reader is referred to that work for a more detailed description of their original model, but we provide an overview here for convenience. The original model code is based on the source code provided accompanying the online dashboard for running simulations using the model of Paltiel et al. ([14]; https://data-viz.it.wisc.edu/covid-19-screening/). The model parameters discussed below are summarized in Table 1.

**Table 1.**
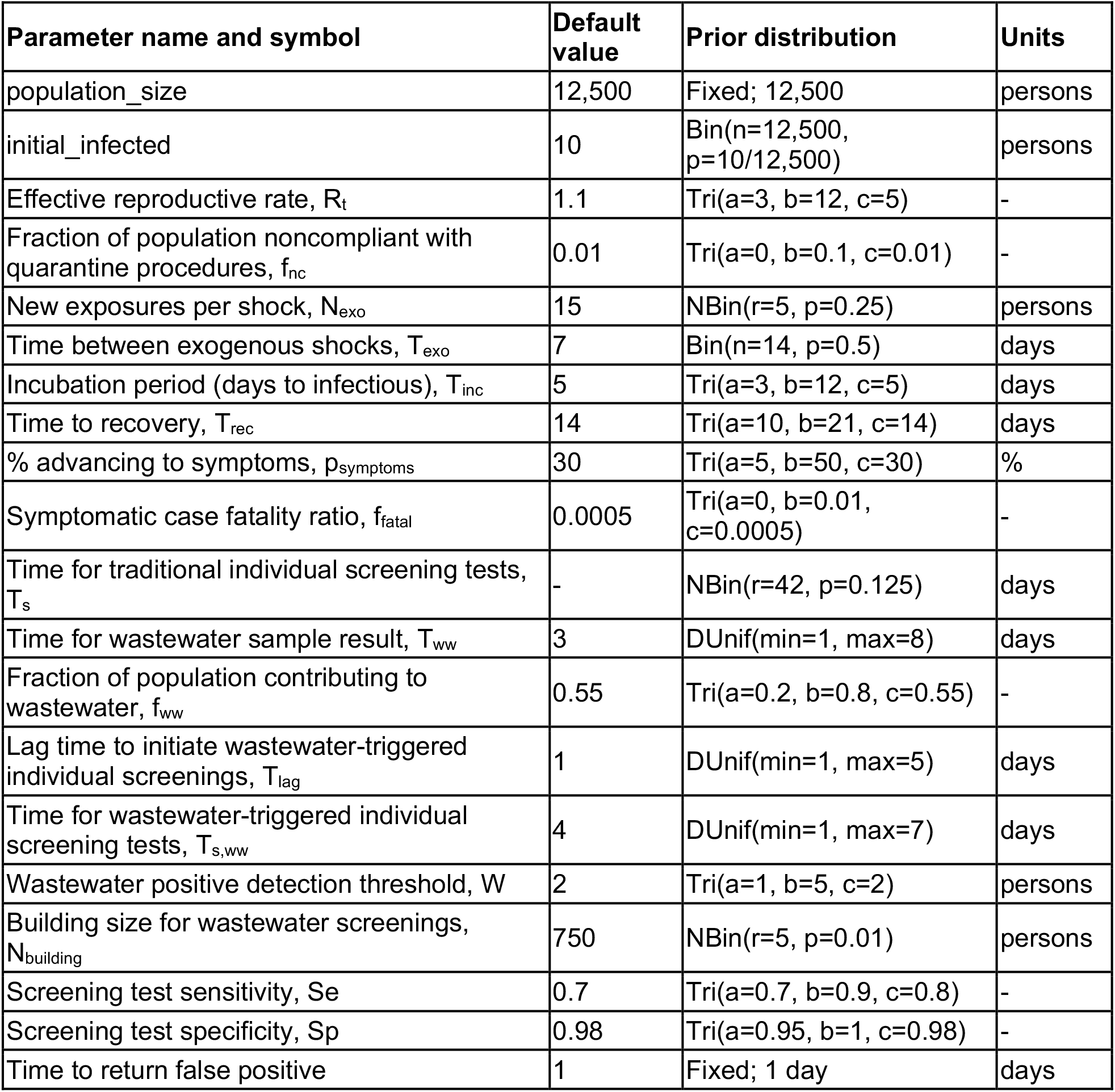
Parameter names, symbols, units, default values for risk analysis, and prior distributions. “Tri” denotes a triangular distribution, which is defined by its range parameters (a and b) and their mode parameter (c). “Bin” denotes a binomial distribution, which is defined by a number of Bernoulli trials (n) and the probability of “success” of one of those trials (p). “NBin” denotes a negative binomial distribution, which is defined by a number of Bernoulli successes (r) and the probability of success (p). “DUnif” and “Unif” denote discrete uniform and continuous uniform distributions, respectively.

The overall campus population is divided into three groups. The first group contains individuals who are circulating on campus and are capable of becoming infected by others, or capable of infecting others. This includes individuals who are uninfected and susceptible (U), those who have been exposed to an infectious individual but are not symptomatic and not infectious (E), and those who have been infected and are asymptomatic but infectious (A). The second group contains individuals who are in quarantine/isolation, including those who have been tested and received either a false or true positive result (FP and TP, respectively) and those who are symptomatic (S). The third group contains individuals who have been removed from the pool of potentially infected/infectious persons, including those who have been infected but have since recovered (R) and those who were infected and later died (D). The model tracks the total number of individuals in each of these model compartments, but not the individuals themselves. Fig 1 provides a schematic of these model components and the exchanges of individuals between them. We assume that the overall size of the campus population is 12,500 individuals, and that initially A(0)=10 of them are asymptomatic and infectious.

**Figure 1.**
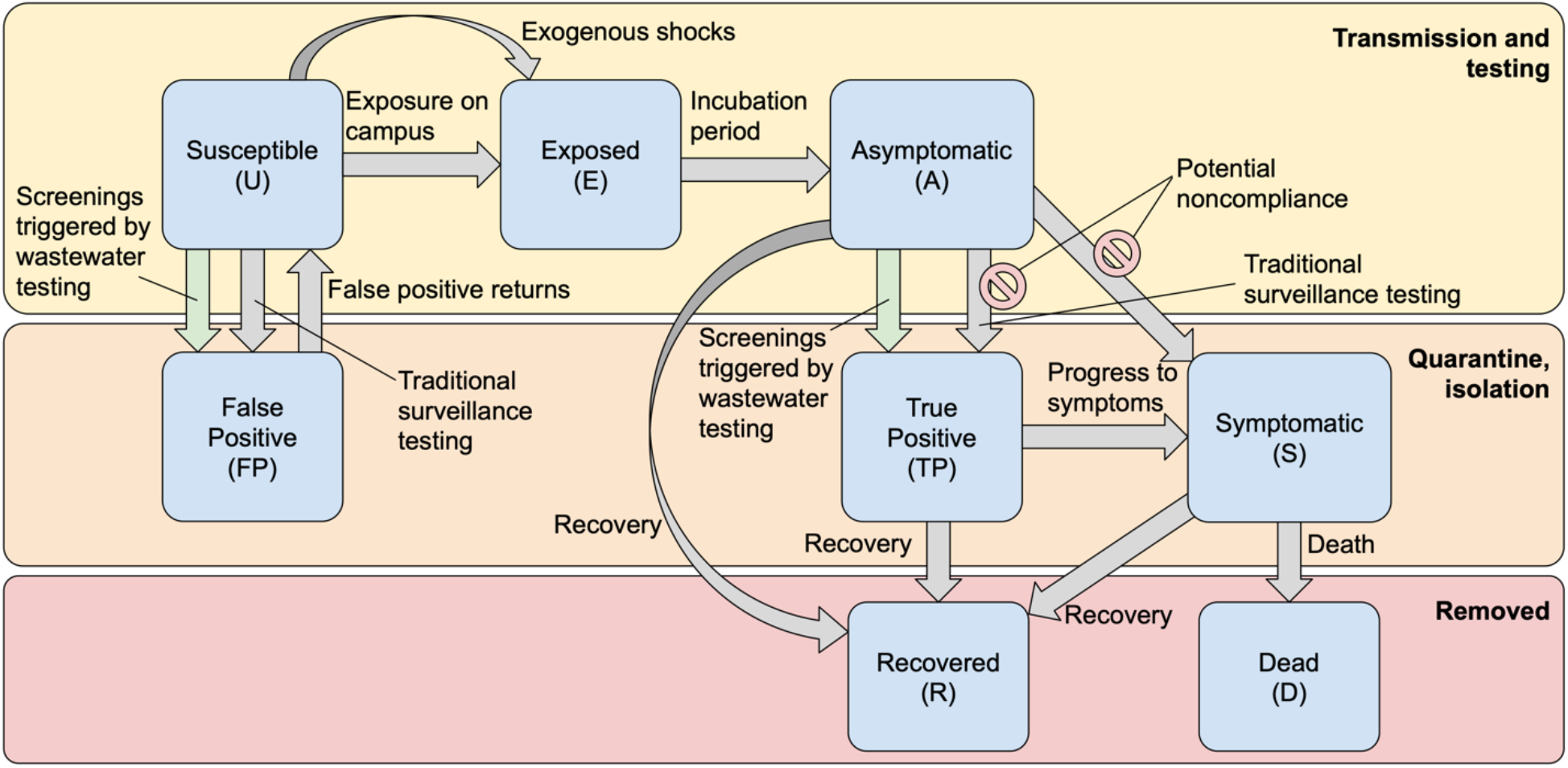
Model schematic with “fluxes” of individuals between model compartments. This includes potential noncompliance with quarantine/isolation procedures and the new wastewater fluxes based on screenings triggered by wastewater-positive results. Noncompliance reduces the flow of Asymptomatic individuals into the True Positive or Symptomatic compartments. Note that Recovered individuals are still circulating on campus, but are assumed to have been removed from the set of individuals who can become infected or transmit the virus.

Susceptible individuals (U) become exposed (E) at a rate determined by the effective viral reproduction rate (R_t_) and the overall prevalence of asymptomatic infectious individuals (A) in the circulating group. We take R_t_ as an uncertain input parameter, with a default value of 1.1, which (as of this writing) is appropriate for Upstate New York where our home institution is located [15,16]. We consider uncertainty in R_t_ and the impacts of incorrect assumptions about the value of this key parameter in the sensitivity and risk analyses described in Sections 2.3 and 2.4. Susceptible individuals (U) also become exposed due to exogenous shocks, such as infections of campus community members by outside individuals or “superspreader” events. The time between exogenous shocks (T_exo_) and number of new exposures from each shock (N_exo_) are both taken as uncertain input parameters (see Table 1). Exposed individuals (E) become infectious and transition into the asymptomatic and infectious (but still circulating on campus) group (A) at a rate governed by the incubation period, T_inc_. We take T_inc_=5 days as a default value [4,17], but also consider uncertainty in this parameter in our subsequent analyses.

We assume that asymptomatic infectious individuals (A) develop symptoms at rate σ (transition to S), and individuals who are infectious recover at a rate ρ (transition to R), regardless of whether or not they exhibit symptoms. Following Paltiel et al. [4], we assume that the percentage of infected individuals that eventually develop symptoms (p_symptoms_) is 30%. Thus, p_symptoms_=0.3=σ/(σ+ρ). Letting T_rec_ be the recovery time from the infection, then ρ=1/T_rec_. We take T_rec_=14 days as a default parameter value, which leads to σ=3/98 days^-1^ as a default. Individuals who are symptomatic perish at rate δ, which depends on the recovery rate (ρ) and the symptomatic case fatality ratio, f_fatal_. We take f_fatal_=0.05% as a default value. We note that values for this parameter are likely to be higher for certain subpopulations of the campus population (for example, groups designated as high-risk by the United States Centers for Disease Control and Prevention (CDC) [18,19]. We assume that recovered individuals (R) are no longer susceptible to infection. As our model simulations are for a single semester (about 14 weeks), this is in line with the current thinking on the length of protection offered by SARS-CoV-2 antibodies [20,21].

Traditional individual surveillance testing is carried out with period T_s_ so that the entire circulating campus population is screening once every T_s_ days. Note that the T_s_ parameter represents how long it would take to test the entire campus population, but this testing is divided up so that some portion occurs in each model time step. The screening test sensitivity (Se, true positive rate) and specificity (Sp, true negative rate) are taken as uncertain input parameters. We take Se=70% and Sp=98% (false negative and false positive rates of 30% and 2%, respectively) as default values, together with the prior distributions described below. Following the original work of Paltiel et al. [4], we assume that false positives are detected via a confirmatory test afterward with 100% specificity. The time to return a false positive to the susceptible circulating group is an input parameter that we take to be 1 day as a default. Also following Paltiel et al. [4], testing of exposed, but not infectious or symptomatic, individuals (E) is assumed to always lead to a negative screening result. This assumption is made in light of the practical difficulty involved in tracking how long each member of the exposed group has spent in that model compartment, as the proportions of false positives and false negatives are affected by the prevalence of viral RNA in the affected individuals. Additionally, as screening tests are carried out, some of the exposed individuals will transition into the asymptomatic (infectious) compartment, where they will potentially be screened as true positives.

In the original formulation of Paltiel et al. [4], it is assumed that individuals who develop symptoms or receive a positive screening test result immediately are moved into isolation/quarantine. We have added a parameter to represent noncompliance with isolation/quarantine procedures. This parameter, f_nc_, is a fraction between 0 and 1 that represents the proportion of the asymptomatic and circulating population that does not go into quarantine even after either developing symptoms or testing positive for SARS-CoV-2. f_nc_ can also represent the event that an individual develops symptoms but those symptoms are so mild that they do not realize they are infected. We take this parameter to be equal to 0 by default. However, in light of recent news of student noncompliance with quarantine/isolation procedures [22–24], it is important to evaluate the impacts of student noncompliance on the efficacy of campus strategies to manage the spread of SARS-CoV-2. We note that f_nc_ only represents noncompliance with quarantine/isolation rules, and does not represent noncompliance with limitations on large gatherings or mask-wearing, for example. Restrictions on gathering sizes and mask mandates are represented in the effective reproductive rate, R_t_.

### 2.2 Wastewater sampling

#### 2.2.1 Overview

We begin with a brief overview of the wastewater module that we have added to the SEIR model of Paltiel et al. [4]. Then, in Section 2.2.2, we provide specific details regarding the implementation in the model structure.

Wastewater from a handful of different sewer sites around campus is tested for signs of SARS-CoV-2 every few days. In the real world, if there are signs that the virus is present in one or more of these wastewater sampling locations, then the relevant wastewater sample is considered to be “positive” and screening tests will be administered to the subpopulations on campus that would have contributed to those wastewater samples. The limit of detection will vary between campuses and the specific wastewater collection and testing systems employed [25]. In our model, a positive wastewater sample is triggered when the number of asymptomatic or exposed (but not yet infectious) individuals who contribute to the wastewater samples exceeds a threshold parameter.

#### 2.2.2 Specific implementation

We assume that a set of wastewater samples is drawn from sewer systems around campus and sent to a laboratory for testing every at regular intervals, T_ww_. We assume that wastewater sample results return from the laboratory every T_ww_ days as well. Thus, at time t=n×T_ww_, the wastewater results from time t=(n-1)×T_ww_ return from the laboratory, where n is a positive integer. As a default parameter value, we take T_ww_=3 days. We assume that a proportion, f_ww_, of the overall population is contributing to wastewater samples, and that the population is well-mixed. This subpopulation consists of students who live on campus, and students, faculty, and staff who work or study on campus. A positive wastewater sample result is returned in the model if the number of asymptomatic and exposed individuals contributing to the wastewater samples at the time that the wastewater sample is drawn exceeds a threshold parameter, W (i.e., wastewater sample is found to indicate the presence of viral RNA). Based on results from University of Arizona [26] and preliminary analyses at our home institution, we take W=2 as a default value. We include uncertainty in W in the sensitivity and risk analyses described in Sections 2.3 and 2.4.

For every college campus that is implementing a wastewater surveillance system, the building(s) that each sewage sample represents will be different. Thus, if there is evidence of viral RNA found in the wastewater sample from a particular sewer draw, it is unknown exactly how many individuals contributed to that particular sample. In light of this uncertainty, we use a parameter, N_building_, as a representative number of individuals that would contribute to an arbitrary wastewater sample. This can represent a single large residence hall or a collection of several smaller buildings. Uncertainty in N_building_ also arises from the fact that different sewers from which a campus might sample will represent (collections of) buildings of different capacities, and multiple sewers can yield positive results simultaneously. We use a default value of N_building_=750 persons. 750 is toward the upper end of the range of building capacities for our home institution. If a positive wastewater signal leads to the testing of, on average, fewer than N_building_ individuals, then choosing higher values for N_building_ reflects the belief that sometimes multiple sewer systems will give positive wastewater results simultaneously.

We assume that the wastewater-triggered individual screening tests begin some time, T_lag_, after receiving the positive wastewater result. By default, we take T_lag_=1 day. There are practical reasons to potentially delay initiating screening tests after a positive wastewater result. For example, a large portion of the exposed individuals who are not yet infectious might go undetected if screening tests are administered before they have transitioned into the asymptomatic (and infectious) model compartment.

Each time that a wastewater sample is returned from testing (i.e., at intervals of T_ww_), the model checks if the wastewater result is positive. If a positive wastewater result occurs and screening tests begin, then we assume that all f_ww_×(E(t)+A(t)) exposed and asymptomatic individuals contributing to wastewater sampling reside within the same representative building (or set of buildings) of size N_building_. It is important to note that this might represent a collection of actual buildings; hence, our choice to use a value for N_building_ that is toward the upper end of the set of values for our campus. Here, t represents the current day in the simulated semester (T_ww_+T_lag_ days since the wastewater sample in question was drawn), E(t) represents the number of exposed individuals at time t, and A(t) represents the number of asymptomatic individuals at time t. We assume that the remaining N_building_ - f_ww_×(E(t)+A(t)) individuals are divided between the susceptible (U(t)) and recovered (R(t)) groups. The relative proportions of this remaining building subpopulation that are susceptible and recovered are the same as those proportions in the general campus population. Following the original implementation of Paltiel et al. [4], we assume that the exposed group (E(t)) will always yield a true negative result when tested for SARS-CoV-2, but they can contribute to a positive wastewater signal [9,25].

The time required to administer tests and receive results for the size N_building_ subpopulation that contributes to the wastewater samples is T_s,ww_. As a default, we assume that all individuals contributing to a positive wastewater result will be screened within T_s,ww_=4 days. Similarly to our modification of the traditional individual surveillance testing, the false and true positive results from the wastewater-based screenings are modulated by a noncompliant proportion parameter, f_nc_. After wastewater-triggered individual screening tests remove detected infections from the wastewater-contributing subpopulation, the relative proportions of asymptomatic individuals in the wastewater-contributing subpopulation can be lower than the proportion among the general campus population. Our assumption of a well-mixed general population means that asymptomatic cases from outside the wastewater-contributing population will redistribute into the wastewater-contributing population. This may be viewed as the advent of a new outbreak in a different set of buildings than the set that was just administered the screening tests. An agent-based model would offer an opportunity to investigate these dynamics further, but is beyond the scope of the present work.

### 2.3 Sensitivity analysis

We use the Sobol’ method for global sensitivity analysis [27] to evaluate the sensitivity of our model to each of its input parameters and their interactions with one another. Global sensitivity analyses are preferable to one-at-a-time sensitivity analyses in the sense that a one-at-a-time analysis risks missing important interactions among uncertain parameters. However, global analyses are more computationally expensive due to the larger number of effects to explore. Here, our simple SEIR model is sufficiently inexpensive that running hundreds of thousands of simulations is possible on a time-scale of hours, so a global sensitivity analysis is feasible.

The Sobol’ method examines how the variance in a model output of interest changes when the model input parameters are all varied simultaneously. Via our Sobol’ analysis, we decompose the variance in the total cumulative number of infections over a 100-day semester into portions attributable to each input parameter, and to each parameter interaction. The portion of variance in the model output that is directly attributable to changes to an individual parameter is that parameter’s first-order sensitivity index. The second-order Sobol’ sensitivity indices are the proportions of variance in the model output that are attributable to pairs of model input parameters that are varied together. Higher-order indices may be computed as well, but are typically not presented due to challenges of visualization and the large number of possible three-parameter combinations (for our model, there are over 800 such triplets). However, we also compute the total sensitivity index for each parameter, which is the proportion of variance attributable to that parameter and all of its interactions with other parameters, including these higher-order interactions.

We assign each model input parameter a marginal prior probability distribution (Table 1 and Supplementary Material). We sample from these prior distributions to create two independent ensembles of 10,000 simulations using a Latin hypercube sampling approach [28]. The Sobol’ method computes the parameters’ sensitivity indices by constructing new simulations by swapping values for each parameter, and combinations of parameters, from one ensemble to the other. For example, the second-order sensitivity to R_t_ and N_exo_ would be computed by swapping all of the values for R_t_ and N_exo_ from the second ensemble into the first, and observing the change in model output variance from when all parameters were from the first ensemble. To estimate all of the first-order, second-order, and total sensitivity indices, a grand total of 380,000 model simulations are required. We use bootstrap resampling with 1,000 replicates to compute 95% confidence intervals for each sensitivity index. We diagnose convergence when the widest confidence interval has a width that is less than 10% of the highest total sensitivity index (e.g., 29). We report only sensitivities that constitute at least 1% of the total variance in the modeled total cumulative number of infections, and whose 95% confidence interval excludes 0.

In the sensitivity analysis, we assume our hypothetical campus pursues a testing strategy that relies primarily on wastewater surveillance, but is complemented with a small amount of traditional individual surveillance testing.

### 2.4 Risk analysis

Campus decision-makers would seek to optimize obvious objectives such as minimizing the total number of infections or minimizing the total number of screening tests needed. Public health mandates may also provide additional objectives that decision-makers aim to satisfy. Here, we consider the ability of our university to maintain fewer than 100 new infections across any 14-day period. This is the threshold beyond which universities in New York State would need to switch to all-online classes for at least two weeks [30]. We note that there are specific 14-day periods in which infection counts are tabulated, but examine the objective of maintaining fewer than 100 new infections across any such period, as this will ensure that the state requirement is met. Additionally, we note that smaller institutions would need to close upon reaching a new infection count equal to 5% of their population, and that local health departments may elect to keep the institution closed for in-person classes longer as the situation demands. For brevity’s sake, for a given model simulation, we denote the maximum number of new infections across any 14-day window as I_max,14_. While a testing strategy (wastewater and/or traditional individual surveillance) might satisfy the I_max,14_<100 objective under nominal conditions (default parameter values), incorporating uncertainty in the model parameters by sampling from their prior distributions will lead to a corresponding distribution of I_max,14_ values for the given testing strategy. We define the reliability of maintaining I_max,14_<100 as the probability Pr(I_max,14_<100). This probability, of course, is conditioned on the testing strategy as well as the underlying parameter prior distributions and other model structural assumptions. With these caveats in mind, we examine which wastewater testing strategies provide the highest reliability of maintaining I_max,14_<100 infections.

Guided by our hypothesis that uncertainty in model input parameters will diminish the ability of our hypothetical university to satisfy its objectives for containment of the SARS-CoV-2 virus on campus, we conduct a Monte Carlo risk analysis. We consider surveillance testing strategies that involve only wastewater surveillance, as traditional surveillance testing (e.g., nasal swabs and/or saliva tests) has been considered extensively elsewhere in the literature [4,5]. We consider testing strategies with a time lag to begin wastewater screenings after finding a positive result in the wastewater samples of T_lag_=1 day. We consider times to complete all wastewater-triggered screening tests, which begin after the 1-day lag, of T_s,ww_=1, 2, …, 7, or 8 days. Preliminary experiments (see Supplemental Material) suggest that the lag time to initiate wastewater screenings did not substantially (>1%) affect the resulting infection rates. This is because eventually, infections are so prevalent on campus that the wastewater-triggered screenings become a nearly continuous process.

We evaluate the sensitivity of these eight testing strategies, corresponding to the eight values of T_s,ww_ above, to our assumptions about three critical parameters: the effective reproductive rate, R_t_, the number of new infections from exogenous sources each week, N_exo_, and the fraction of individuals who are not compliant with quarantine/isolation procedures, f_nc_. Specifically, we consider a control scenario in which we assume that R_t_=1.1, N_exo_=15 on a weekly time-scale (T_exo_=7 days), and f_nc_=0.01 (1%). In the risk analysis, we suppose that the hypothetical campus uses only wastewater-triggered individual testing and does not conduct any additional traditional individual testing (e.g., nasal swab or saliva). We let T_s,ww_ vary at its values stated above, we keep the parameters T_lag_, R_t_, N_exo_, T_exo_, and f_nc_ fixed, and we sample all of the other model parameters from their prior distributions (Table 1). We assume that the total campus population is fixed (12,500 individuals), and do not consider uncertainty in the time to return false positive results (1 day). We estimate the reliability of maintaining I_max,14_<100, Pr(I_max,14_<100), as the proportion of simulations in which the maximum number of new infections across any 14-day window does not exceed 100. For each testing strategy and combination of R_t_, N_exo_, and f_nc_, we generate 3,000 sets of the other parameters from their marginal prior distributions using a Latin Hypercube sampling approach [28]. We find that at least about 2,000 samples are required in order to stabilize our estimates of Pr(I_max,14_<100), subject to the uncertainties in the other model parameters (see Supplemental Material).

We then consider how “breaking” our assumptions about the parameters R_t_, N_exo_, and f_nc_ affects the reliability Pr(I_max,14_<100). We construct seven additional ensembles by increasing R_t_ to 1.5, increasing the number of new exposures for each weekly shock to 30, and increasing the fraction of noncompliant individuals to f_nc_=0.1, as well as all combinations of these increases. By observing the decrease in Pr(I_max,14_<100), we quantify the fragility of each testing time-scale to broken assumptions about the rate of viral transmission on campus, the influence of infections of campus community members by outside sources, and noncompliance by students.

Importantly, in the simulations that we present here, there are two total infection counts. First, there is the number of true infections, including asymptomatic cases (A(t)), symptomatic (S(t)), and true positive cases (TP(t)). The number of true infections is generally unknown in real life. By contrast, our model also computes the number of perceived - or detected - infections.

The number of perceived infections includes symptomatic cases (S(t)), true positives (TP(t)) and false positives (FP(t)), but misses asymptomatic cases. We thus present two sets of reliabilities: one using the true number of infections and one using the perceived infections only. This distinction, and the fact that in the “model world” the true number of infections is known, enables us to characterize how a lack of timely testing can obscure the true infection count. So, we hypothesize that as the testing time T_s,ww_ increases, the perceived reliability will increase, while the true reliability will decrease. However, we hypothesize that beyond a certain length of time for testing, the perceived reliability will begin to decrease because the number of symptomatic cases becomes overwhelming.

## 3 Results

### 3.1 Comparison with traditional individual surveillance testing

Before embarking on our Monte Carlo sensitivity and risk analysis, we first examine individual simulations under the control scenario as described in Sec. 2.4. We use the default parameter values from Table 1, including weekly 15-infection shocks (T_exo_=7 days, N_exo_=15), R_t_=1.1, and f_nc_=0.01 noncompliance. Using wastewater surveillance with T_lag_=1 day and T_s,ww_=4 days leads to true numbers of infections that are quite similar to the infection count under a traditional surveillance testing regimen with T_s_=7 days (Fig 2A, solid lines). The total cumulative numbers of infections under the wastewater and traditional surveillance approaches were 410 and 399 infections, respectively, at the end of the 100-day model simulations. This difference constitutes an increase of 2.8% of the number of true infections under the traditional screening approach. Under the control scenario, the projected numbers of deaths for the semester are 0.043 and 0.042 for the wastewater and traditional screening cases, respectively. These differences increase slightly in the moderate (R_t_=1.5, N_exo_=20, and f_nc_=0.05; Fig 2C) and severe (R_t_=2, N_exo_=30, and f_nc_=0.1; Fig 2E) scenarios. In the moderate scenario, there are 860 total infections (0.082 deaths) projected under the wastewater surveillance program and 828 infections (0.079 deaths) under the traditional surveillance testing program. In the severe scenario, there are 3,825 infections (0.30 deaths) and 3,655 infections (0.29 deaths) using the wastewater and traditional surveillance approaches, respectively. The numbers of true infections when using wastewater surveillance constitute increases of 3.9% and 4.7% over the 1-week traditional surveillance plan in the moderate and severe risk cases, respectively.

**Figure 2.**
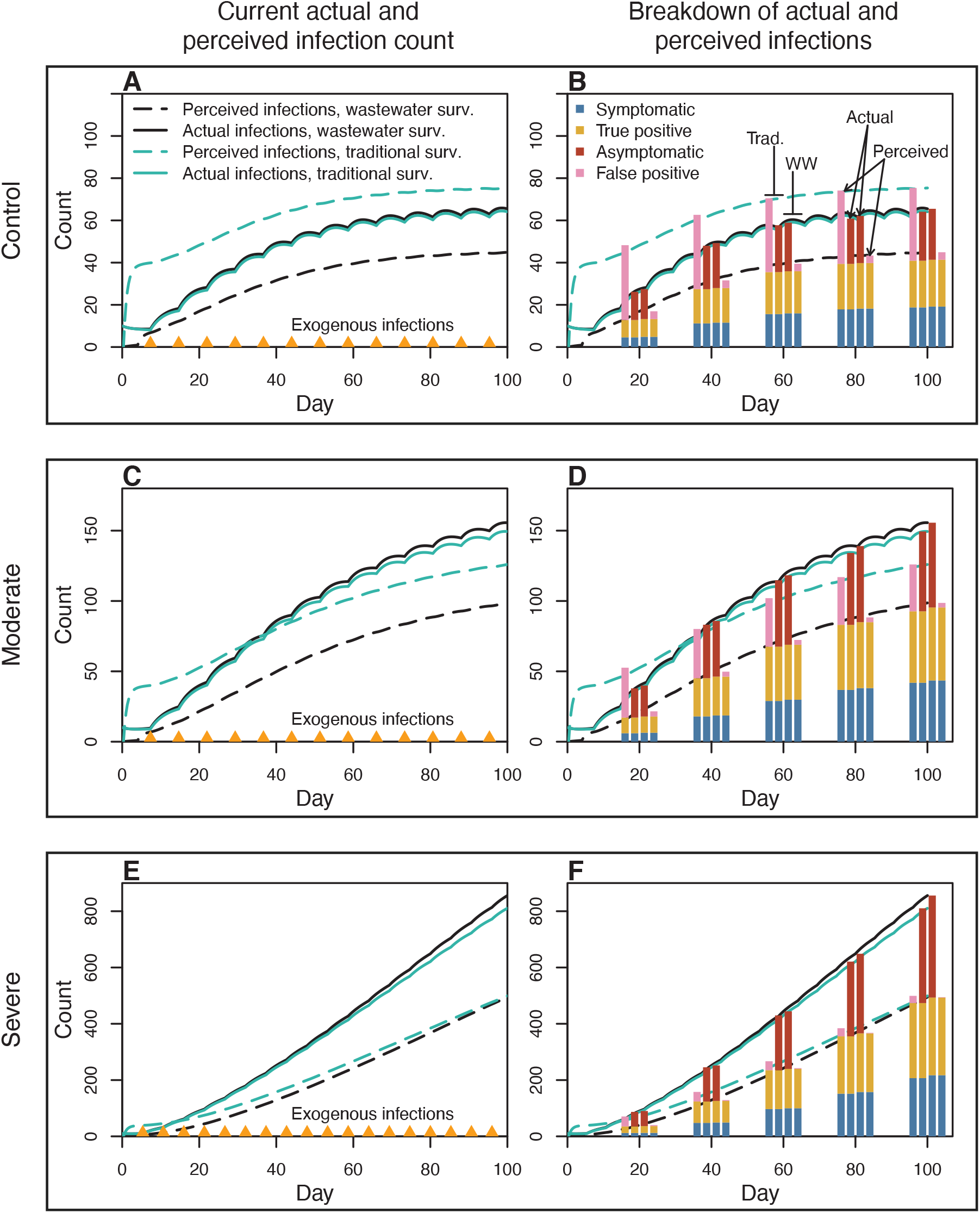
Wastewater surveillance (black lines) underestimates the true number of infections (solid lines), while traditional individual surveillance (green lines) tends to overestimate it for low-risk scenarios, then underestimate it for high-risk scenarios. Shown are the numbers of current infections at each time throughout the semester (A, C, and E) and the breakdown of those infections among the asymptomatic, symptomatic, true positive, and false positive model compartments (B, D, and F) at 20-day intervals. In the right column, the middle bars in each group represent the actual numbers of infections and the outside bars represent the perceived infections. The first two bars in each group represent traditional individual surveillance and the last two bars represent wastewater surveillance. The nominal scenario (first row) uses the parameter values from Table 1, including R_t_=1.1, N_exo_=15, T_exo_=7 days, and f_nc_=0.01. The moderate risk scenario (second row) increases R_t_ to 1.5, increases N_exo_ to 20, and f_nc_=0.05, while all other parameters are the same as in the control scenario. The severe risk scenario (bottom row) uses R_t_=2, N_exo_=30, T_exo_=5 days, and f_nc_=0.1. The occurrences of exogenous infections (of size N_exo_) are denoted with orange triangles in the first column. All of the wastewater screening cases use T_lag_=1 day and T_s,ww_=4 days, and all of the traditional individual screening cases use T_s_=7 days (weekly screenings).

While the numbers of true infections are similar, the numbers of perceived infections are quite different. Under the control scenario, our model predicts that using wastewater surveillance leads to a dramatic underestimation of the number of true infections: at the end of the semester, there are about 45 perceived infections and 66 true infections. This is the result of wastewater surveillance testing a sicker subpopulation resulting in many fewer false positive individuals than traditional surveillance testing (Fig 2B). Traditional surveillance overestimates the number of true infections under the control scenario (Fig 2A, green lines), but this overestimation is the result of a larger portion of false positive individuals and not from detecting more asymptomatic cases (Fig 2B). The sizable number of false positive cases ensnared by the traditional individual surveillance testing strategy can be understood by a quick heuristic calculation: out of a campus population of 12,500 individuals, 12,500/7≅1,786 of them are administered a test on the first day of the simulated semester. With a nominal false positive rate of 2% (1-specificity), we expect to see about 1,786×0.02≅36 false positive cases. This roughly matches the false positive census by day 20 seen in Fig 2B. However, using traditional surveillance in the moderate scenario initially overestimates the number of true infections, then at day 37, the number of true infections becomes larger than the number of perceived infections (Fig 2C, green lines), which thus underestimate the number of true infections after day 37. For the severe risk scenario, this switch occurs after only 15 days (Fig 2E). One of the reasons a campus might pursue a traditional surveillance testing approach is to err on the side of a Type 1 error (false positive), when the potential drawbacks to missing positive COVID-19 cases are high (e.g., an outbreak). These results suggest that the utility of this traditional approach relies on the testing strategy being sufficiently aggressive as to match the local viral prevalence and spread.

The model prediction that wastewater surveillance does not result in a high portion of false positives is not surprising. The structure of our model assumes only a certain fraction of the campus population contributes to the wastewater samples (f_ww_=0.55 is the nominal parameter value). Indeed, not all members of the campus community will be equally represented in wastewater samples. Wastewater surveillance enables the model to identify a subpopulation of sicker individuals and focus screening tests on this subpopulation. Of course, this also means that many individuals will not be within the group that is administered wastewater-triggered screening tests. In practice, wastewater sampling should be used in tandem with some traditional individual surveillance testing to address this issue. However, even in the moderate and severe scenarios, the number of overall infections is never more than 5% higher when using wastewater-based surveillance testing as opposed to traditional individual surveillance testing (Fig 2C and 2E). Thus, it appears that for the scenarios considered here, the infections that wastewater sampling misses do not lead to large outbreaks of infections.

### 3.2 Sensitivity analysis

For the set of model input parameters and prior distributions chosen to reflect conditions with low rates of viral transmission and exogenous exposures, we find that nine parameters, out of 18 total, contribute significantly to variance in the overall number of infections (Fig 3). The criteria for “significance” is for the sensitivity index to be at least 1% of the total variance in estimated cumulative infections, and for the 95% confidence interval to exclude 0. The nine sensitive parameters, in order of their first-order sensitivity indices, are: the effective reproductive rate (R_t_, 35%), the number of new exposures per exogenous shock (N_exo_, 12%), the frequency of exogenous shocks (T_exo_, 6%), the fraction of the population contributing to wastewater samples (f_ww_, 4%), the amount of time between wastewater sample results return (T_ww_, 4%), the amount of time for an infected individual to recover (T_rec_, 4%), the fraction of infected individuals who exhibit symptoms (p_symptoms_, 3%), the amount of time for an exposed individual to become infectious (T_inc_, 3%), and the amount of time required to complete screening tests after a positive wastewater result (T_s,ww_, 2%). We find a total sensitivity to R_t_ and its interactions with other parameters of 63%. This highlights the importance of practical measures to reduce R_t_, including reduced classroom and residence hall capacities, limits on large gatherings, and mask mandates, for example.

**Figure 3.**
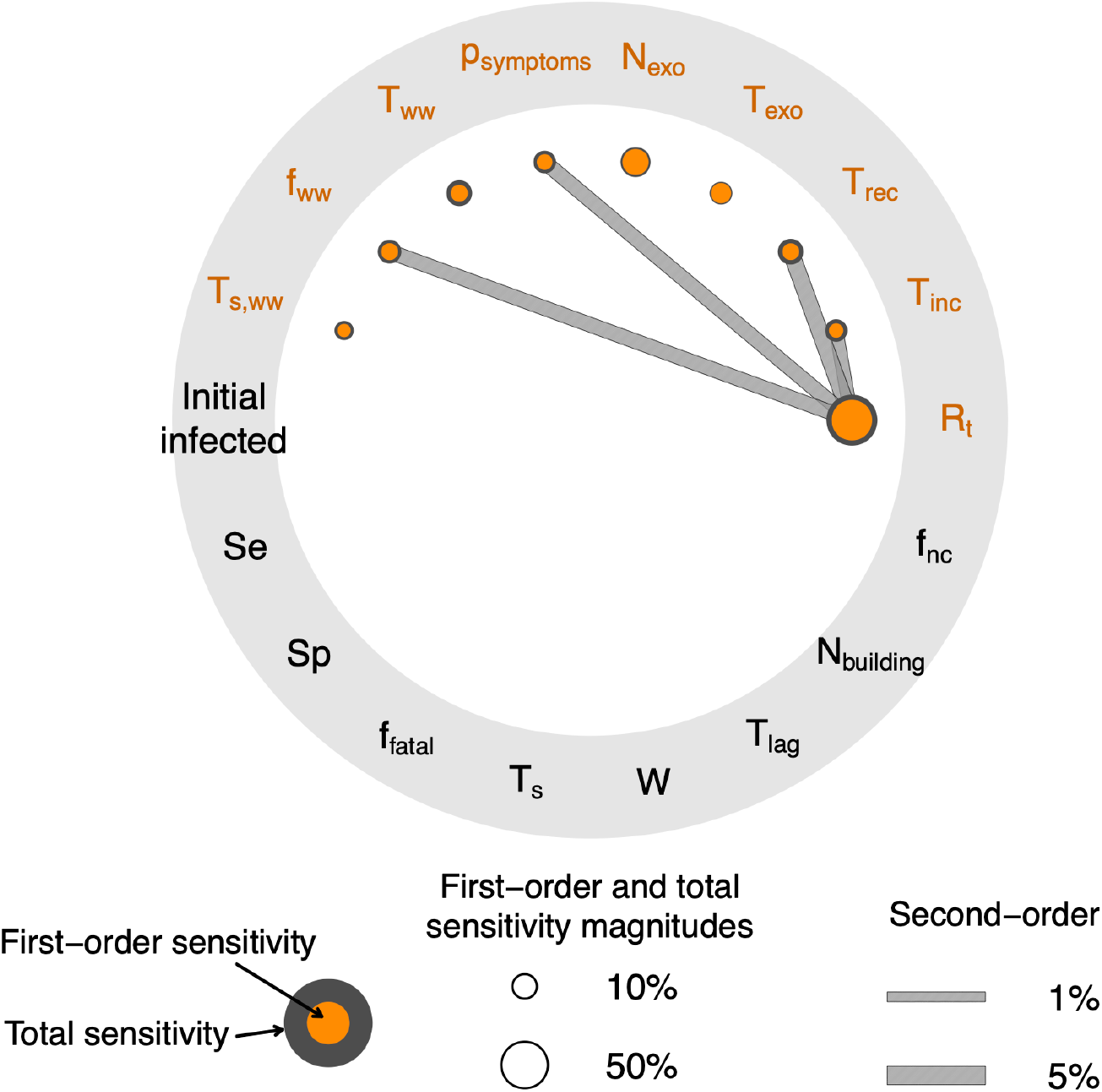
To what degree do the individual and combined parameter uncertainties and decision levers influence the overall cumulative number of infections over the course of the simulated semester? This Sobol’ radial convergence diagram depicts the decomposition of variance in the total number of infections among the uncertain input parameters. Input parameter prior distributions have been chosen to represent the conditions and characteristics of the authors’ home institution, which corresponds to a relatively low value of R_t_, about 10-20 exogenous infections per week, and low rates of noncompliance. Filled orange nodes represent first-order sensitivity indices (direct parameter influences); the concentric gray nodes represent total sensitivity indices (the given parameter’s influence in combination with all other parameters); and filled gray bars represent second-order sensitivity indices for the interaction between the given pair of parameters. Parameters whose names or symbols are orange are the parameters to which the model is sensitive (in the upper hemisphere of the diagram); parameters in black (lower hemisphere) are those to which the model does not display a significant sensitivity.

The fact that two out of the top three parameters to which the total infection count is most sensitive are related to infections of campus community members through interactions with external individuals (e.g., social events and interactions off-campus) highlights the importance of campus and municipal public health measures to limit the spread of the virus. This result also illustrates the practical impacts of members of the campus community making (or not making) safe and responsible choices. We emphasize that these results employ prior distributions for the model input parameters that have been chosen to reflect the conditions and characteristics of our home institution in Upstate New York. We include analogous sensitivity analyses for hypothetical scenarios at a larger university situated within a community with higher rates of transmission and a smaller college with a much lower student population and less active interactions with its surrounding community (see Supplemental Material).

### 3.3 Risk analysis

We now turn to the question: How well can different wastewater surveillance strategies be expected to help maintain low rates of infection on campus? In light of the model’s sensitivities to R_t_ and N_exo_, we examine the fragility of different wastewater surveillance strategies using hypothetical scenarios in which the values for those parameters, and the noncompliance parameter, f_nc_, are more severe than the default values from Table 1. We use a set of eight ensembles to examine the reliability of maintaining fewer than 100 infections during any 14-day period (I_max,14_<100) under different policies for wastewater-triggered surveillance testing (Fig 4). All scenarios use a lag time to initiate screenings of T_lag_=1 day. R_t_, N_exo_, and f_nc_ are held fixed at either their nominal values (R_t_=1.1, N_exo_=15 new infections per week, and f_nc_=0.01 (1%) noncompliance) or their high-risk values (moderate/severe cases: R_t_=1.5, N_exo_=30, and f_nc_=0.1). We assume that no additional traditional individual screening tests beyond those triggered by wastewater sampling are performed, and all other model parameters are sampled from their prior distributions.

**Figure 4.**
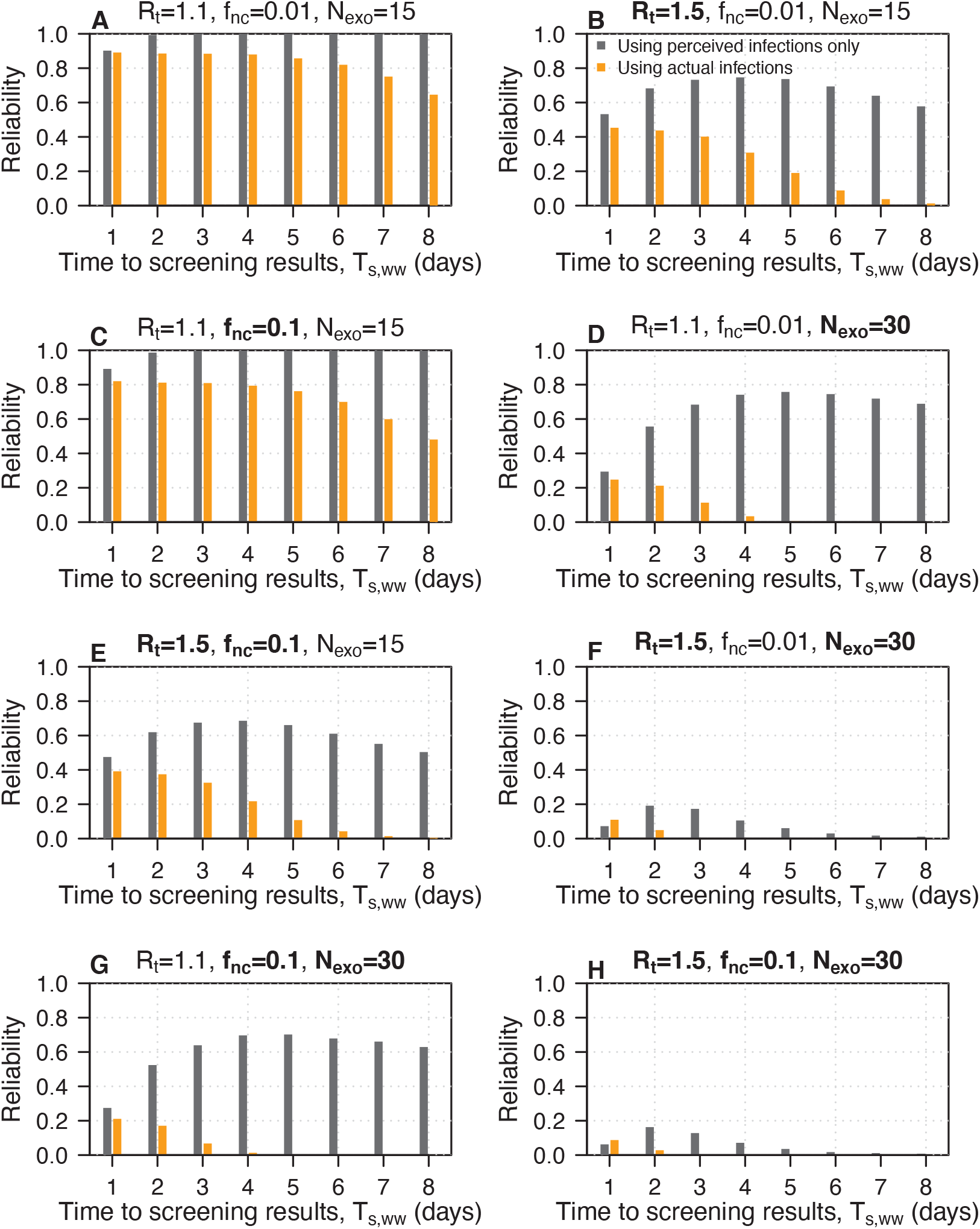
Slower response times to complete screening tests based on wastewater sampling can lead to the perception of safety based on a high reliability of maintaining I_max,14_<100 (gray bars) in contrast to a lower true reliability based on the actual numbers of infections (orange bars). Shown on the vertical axis is the reliability, Pr(I_max,14_<100) and on the horizontal axis are the times required to complete subpopulation screening results after a one-day lag to initiate screening tests. The left bars give the reliability using the actual infection counts and the right bars give the reliability using only the perceived infections, which would hypothetically be known. The panel titles give the values for the reproductive rate (R_t_), the noncompliant fraction (f_nc_), and the number of new exogenous infections each week (N_exo_). Bold-face parameters in the panel titles denote parameters that have taken on values corresponding to higher-risk scenarios. All other parameters are sampled from their prior distributions given in Table 1.

Under nominal conditions, the reliability is about 89% when screenings are completed within 1 day (T_s,ww_ =1), excluding the 1-day lag time to initiate the screenings (Fig 4A). As the time to complete the screening tests increases, the *perceived* reliability increases to 100% (Fig 4A, gray shading) while the *actual* reliability decreases to about 65% with a screening time of 8 days (orange shading). That this discrepancy between the perceived and actual reliability as screening time increases can be seen in all of the higher-risk scenarios as well (Fig 4B-H). This demonstrates how failing to react in a timely fashion to signs of viral RNA can lead to the perception of safety, while in reality the infection count is growing. Additionally, even though both the control scenario and the high-noncompliance scenario yield perceived reliabilities of 100% for screening times longer than 3 days (Fig 4A, C), higher rates of noncompliance lead to about 20-30 additional infections beyond the control scenario (see Supplemental Material).

When R_t_ or N_exo_ are increased to their high-risk values, the actual reliability of maintaining I_max,14_<100 is never greater than 50% (Fig 4B, D-H); the perceived reliability can be up to 76%, with a screening time of 5 days. This reveals the fragility of a surveillance testing strategy that relies on wastewater sampling. Scenarios in which both R_t_=1.5 and N_exo_=30 lead to outcomes in which even the perceived reliabilities do not exceed 20% (Fig 4F, H). For these scenarios, the actual reliability is 0% if the time to receive screening results is longer than 2 days. This highlights the compound hazard associated with multiple drivers of risk on college campuses. Holiday parties, reuniting with friends after winter break, spending more time indoors during winter, and (at some universities) spring break can all serve to increase both R_t_ and N_exo_. This means that these high-risk scenarios must be considered as possible, if not probable, real-world cases as colleges plan for their spring semesters.

We find that across all scenarios, completing the wastewater-triggered screening tests within 1 day of initiating the tests maximizes the actual reliability of maintaining I_max,14_<100. However, in addition to improving student health conditions by maximizing the actual reliability for I_max,14_<100, campus decision-makers in our hypothetical situation would want to also maximize the perceived reliability (based on the perceived number of infections). We emphasize that in practice, the perceived number of infections is the only information known to decision-makers; the actual number of infections is information only known to us here in our model experimental setting. In all scenarios except those with higher R_t_ combined with higher N_exo_, the perceived reliability is maximized by using a screening time of 4-5 days (Fig 4A-E, G). In the two scenarios with both higher R_t_ and higher N_exo_, the perceived reliability is maximized by using a screening time of 2 days (Fig 4F, H). A time to receive screening results of 2 days appears to be a suitable compromise between (i) guarding against high-risk scenarios (i.e., higher R_t_ and N_exo_ simultaneously) and (ii) balancing minimizing true infections against the practical concern of minimizing the number of perceived (detected) infections.

## 4 Discussion

We have tuned a SEIR compartment model to represent scenarios of wastewater screening for SARS-CoV-2 at a hypothetical university of 12,500 students during a time period in which the area is experiencing relatively low viral presence and transmission, as well as low exposure of campus community members from the surrounding community. By developing and implementing a model component for wastewater surveillance, we find that wastewater surveillance testing can be an effective strategy to maintain a similar number of overall infections on campus to traditional individual surveillance testing (Fig 2). This result is conditioned on the assumed values and prior distributions for our parameters, which have been selected to represent situations that might plausibly face our home institution, and is conditioned on the campus health decision-makers responding to signs of infection in a timely manner.

The discrepancy between the true infection count and the number of infections that campus health officials would know about (the perceived number of infections) is striking (Figs 2 and 4). Guarding against the worst-case scenarios that include higher reproductive rate, R_t_, simultaneous with higher exogenous infections, N_exo_, suggests that a time to screening results of T_s,ww_=2 days (plus a 1-day lag to initiate the screenings) is optimal for maximizing the perceived reliability of maintaining I_max,14_<100 (Fig 4F, H). However, in the other higher-risk scenarios, the perceived reliability is maximized by taking T_s,ww_=4 or 5 days (Fig 4A-E, G). Across all scenarios, the actual reliability is maximized by taking T_s,ww_=1 day, the fastest time-to-results considered here. Thus, we suggest that a time of T_s,ww_=2 days to complete wastewater-triggered screening tests balances decision-makers’ desire to avoid a strategy that is fragile against higher-than-nominal risk factors, to minimize the overall number of infections, and the practical importance of maintaining fewer than 100 perceived infections across any 14-day period (Fig 4).

Exogenous infections and R_t_ are the parameters with the highest direct influence on overall number of infections (Fig 3). These results reflect the importance of taking practical steps to mitigate viral spread on campus and within the local community. Additionally, the fact that R_t_ and the frequency and rate of exogenous infections have a stronger influence on overall infection count than the characteristics of the surveillance testing protocol (wastewater or traditional) is likely attributable to the relatively low prevalence and spread of virus in our main test case. In a supplementary experiment, we change the parameters’ prior distributions to represent the case of a large university (20,000 individuals) in an area with higher rates of transmission and exogenous shocks and a small college (3,000 individuals) in an area with low transmission and exogenous shocks (see Supplementary Material). In the large university case, we find that the characteristics of the surveillance testing protocol become more important than the frequency and rate of exogenous infections. The results for the small college case more closely resemble our results for the hypothetical campus of Sec 3.2, with the exception that exogenous infections contribute a relatively smaller portion to the variability in overall numbers of infections.

While our sensitivity analysis and preliminary risk analysis (see Supplementary Material) may appear to indicate that the lag time to initiate screening tests based on a positive wastewater return is inconsequential for improving the reliability, the total number of infections suffers with greater lag times. Furthermore, in many simulations, the prevalence of the virus is high enough that once wastewater-based screening tests are initiated, they continue until the end of the semester (Fig 1). Thus, initiating screening tests within one day, in response to a positive wastewater signal, appears to be the best option to mitigate higher numbers of subsequent infections.

We find that wastewater sampling leads to 410 infections over the course of the entire 100-day semester in the control (low) forcing scenario, in which R_t_=1.1, N_exo_=15 new exogenous exposures each week, and f_nc_=0.01. In the control scenario, the traditional individual surveillance testing approach leads to 399 total infections. Thus, the 11 additional infections under the wastewater surveillance plan represents an increase of about 2.8%. This difference is larger in the moderate (3.9%) and severe (4.7%) forcing scenarios, but remains within 5%. In practice, deploying a blend of wastewater and traditional individual screening approaches would mitigate the effects of this potential issue. Given these general similarities in outcomes, our results suggest that, under nominal or moderate risk conditions, wastewater surveillance is a viable and noninvasive option to monitor the campus population for signs of infection.

Our risk analysis results (Sec 3.3) suggest that perceived reliability - which is the only reliability that will be known in practice - will generally be higher than the actual reliability of maintaining I_max,14_<100. This is driven by the result that wastewater sampling is targeted to a portion of asymptomatic cases (Fig 2). Thus, in practice, we expect that a wastewater sampling plan may appear to keep the overall infection count low, but asymptomatic cases will go unnoticed. By contrast, traditional individual screening tests appear to overestimate the actual number of infections for low- or moderate-risk scenarios (Fig 2A, C). These results have profound practical implications. This result implies that universities that exceed the New York State 100-infection limit and were using a traditional individual surveillance testing approach may not have actually exceeded the 100-infection threshold due to a prevalence of false positive results. On the other hand, universities that employ a wastewater surveillance approach may exceed the 100-infection threshold even if they have an apparent 14-day running total infection count below 100 infections.

Consistent with the results of Paltiel et al. [4], through our global sensitivity analysis, we find that the sensitivity and specificity of the screening tests do not play a central role in contributing to the uncertainty in the overall number of infections. This is, of course, conditioned on the prior distributions that we have chosen for the sensitivity and specificity parameters, which exclude sensitivities less than 70% and specificities less than 95%. As new testing methods become available, it may be necessary to update these prior distributions. Uncertainties in the testing sensitivity and specificity are dwarfed by the uncertainties in on-the-ground conditions and campus health response, including the effective reproductive rate (R_t_) and the frequency and rate of infections of campus community members from off-campus interactions (Fig 3). Additionally, we have chosen the prior distributions for all of the model parameters in order to best represent the conditions facing our home institution in New York State. The choices for the specific forms and parameterizations of these distributions involves many subjective choices, which offer opportunities for further exploration. For example, the times required to carry out surveillance tests (T_s_ and T_s,ww_) could be reparameterized as rate parameters and assigned gamma distributions as their conjugate priors. Here, we retain the time lengths as parameters to reflect the fact that these time-scales are the decision levers that would be directly manipulated by campus decision-makers by prescribing, for example, weekly surveillance testing.

There are, of course, practical steps that a university may take to enhance the efficacy of a wastewater screening protocol beyond what we have illustrated here in our stylized model university. For example, after a wastewater sample result shows signs of infection, our model assumes that all asymptomatic and exposed individuals who triggered the positive result are within an identifiable group of size N_building_, but this group still interacts with other members of the campus community as wastewater-triggered screening tests are administered. Spread of virus may be further mitigated by quarantining the affected building(s). Additionally, our risk analysis focuses on examining how wastewater testing alone can be an effective tool to keep overall infection numbers low. Our results from Sec 3.1 indicate that the reliability of maintaining I_max,14_<100 infections can likely be improved by complementing wastewater screening with traditional individual surveillance testing.

As with any model, the coarse nature of our model and the original model of Paltiel et al. [4] means that the results should not be taken as a prediction of specific future behavior or infection rates. Rather, these models serve as a tool to evaluate the efficacy of various policy decisions to mitigate the spread of COVID-19 on college campuses, and the impacts of uncertainties on infection rates. Our model for wastewater surveillance assumes that some constant fraction, f_ww_, of individuals in the exposed and asymptomatic compartments can contribute to a positive wastewater signal. However, this is an approximation of the reality that the levels of viral shedding in wastewater contributions varies with time during an infection. Additionally, time-variability in the reproductive rate, R_t_, may become important as the Northern Hemisphere enters winter and, in cold and rainy areas, people will spend more time indoors. Specifically, previous work has estimated a winter R_t_ of about 2.2, in contrast to a summer R_t_ of 1.3 [31]. Our risk analysis indicates that such an increase of R_t_ would dramatically diminish the ability of universities to maintain low infection rates (Fig 4B). Taken together, these findings underscore the importance of mask and physical distancing mandates, moratoriums on large gatherings, reduced classroom capacities, and striking a careful balance between in-person and online learning modalities, which can reduce the number of students and faculty circulating on campus at any given time. This balance will of course be different for different campuses, and should be driven by science and evidence that is specific to the unique conditions faced by each university. Decision-makers must carefully consider how the distribution of class modalities affects the numbers of contacts among students each day and how, in turn, this affects transmission rates on college campuses.

## 5 Conclusions

We have presented a model to represent wastewater screening and incorporated it into an existing SEIR compartment model [4] to track the spread of an infection within a college campus community. We find that wastewater surveillance is an effective approach to detect and remove infected individuals from circulating among the campus community. Under nominal or moderate-risk conditions, the model predicts that wastewater surveillance can maintain an overall number of infections similar to the performance of weekly traditional surveillance testing, while also dramatically reducing the number of false positive cases (Fig 2). A period of one day to receive a positive wastewater result, interpret it, and initiate a plan for screening tests, combined with a period of 2 days to complete the screening tests, appears to form the most robust strategy to maintain low infection rates. However, complementing a wastewater surveillance program with conventional surveillance testing via nasal swabs would offer improvements. These results have practical importance for developing strategies to mitigate the spread of COVID-19 on college campuses.

## Data Availability

All model codes, analysis codes, input files, and output data sets are freely available at https://github.com/tonyewong/SEIR-WW under the MIT open-source license.

## 6 Acknowledgements

We thank Enid Cardinal, Wendy Gelbard and Lindsay Phillips for informative conversations about the surveillance testing plans for the Rochester Institute of Technology. We thank A. David Paltiel for assistance interpreting the original SEIR model. We thank Srikanth Aravamuthan, Sean Kent, Steve Goldstein, and Brian Yandell for sharing the source code for the original SEIR model and providing the SEIR model dashboard (https://data-viz.it.wisc.edu/covid-19-screening/). Any views expressed in this work are solely those of the authors and do not necessarily reflect the views of our home institution.

## 8 Author Contributions

GT and TW conceived the study. GT, DR and NB wrote the original exploratory model for the RIT test case. TW wrote the modified model for wastewater sampling, ran the simulations, and wrote the first draft of the manuscript. All authors analyzed the model output, developed the figures, and contributed to the final versions of the model and the manuscript.

