## Supplementary Material for "Evaluating the Sensitivity of SARS-CoV-2 Infection Rates on College Campuses to Wastewater Surveillance"

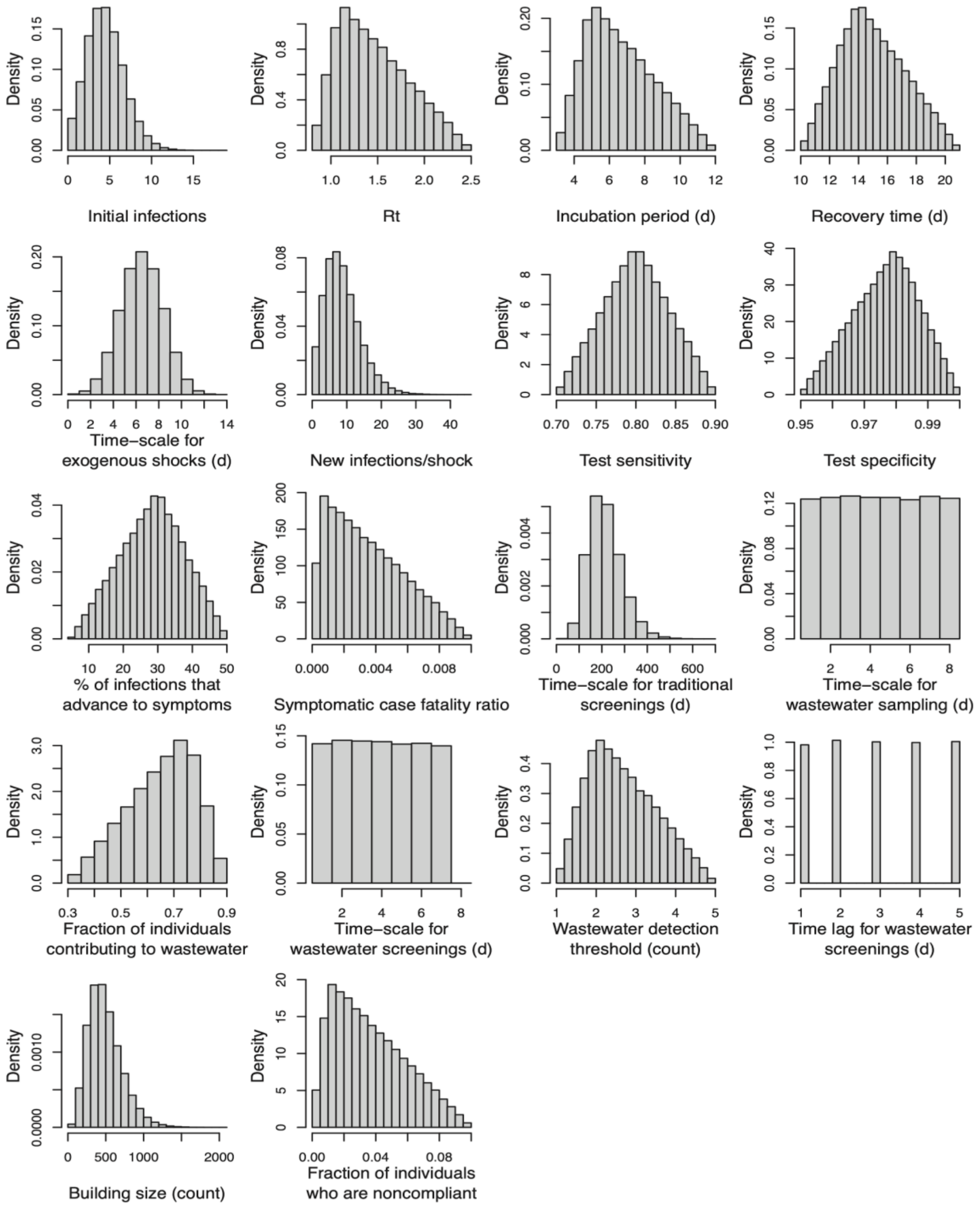


**Figure S1.** Parameter prior distributions for the control scenario (see Table 1). Histograms show 100,000 samples from each parameter's marginal prior distribution.

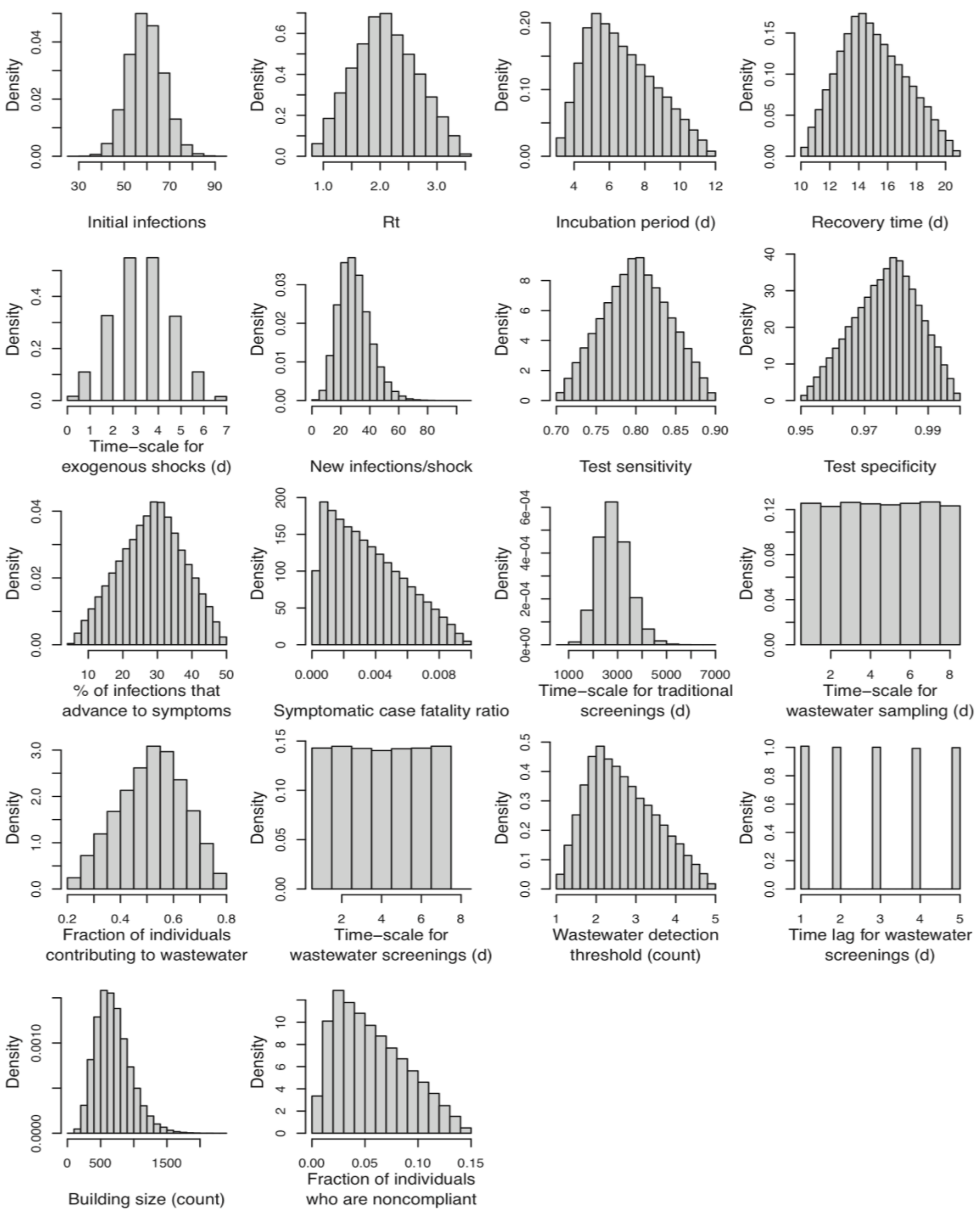


**Figure S2.** Parameter prior distributions for the high-risk scenario. Histograms show 100,000 samples from each parameter's marginal prior distribution.

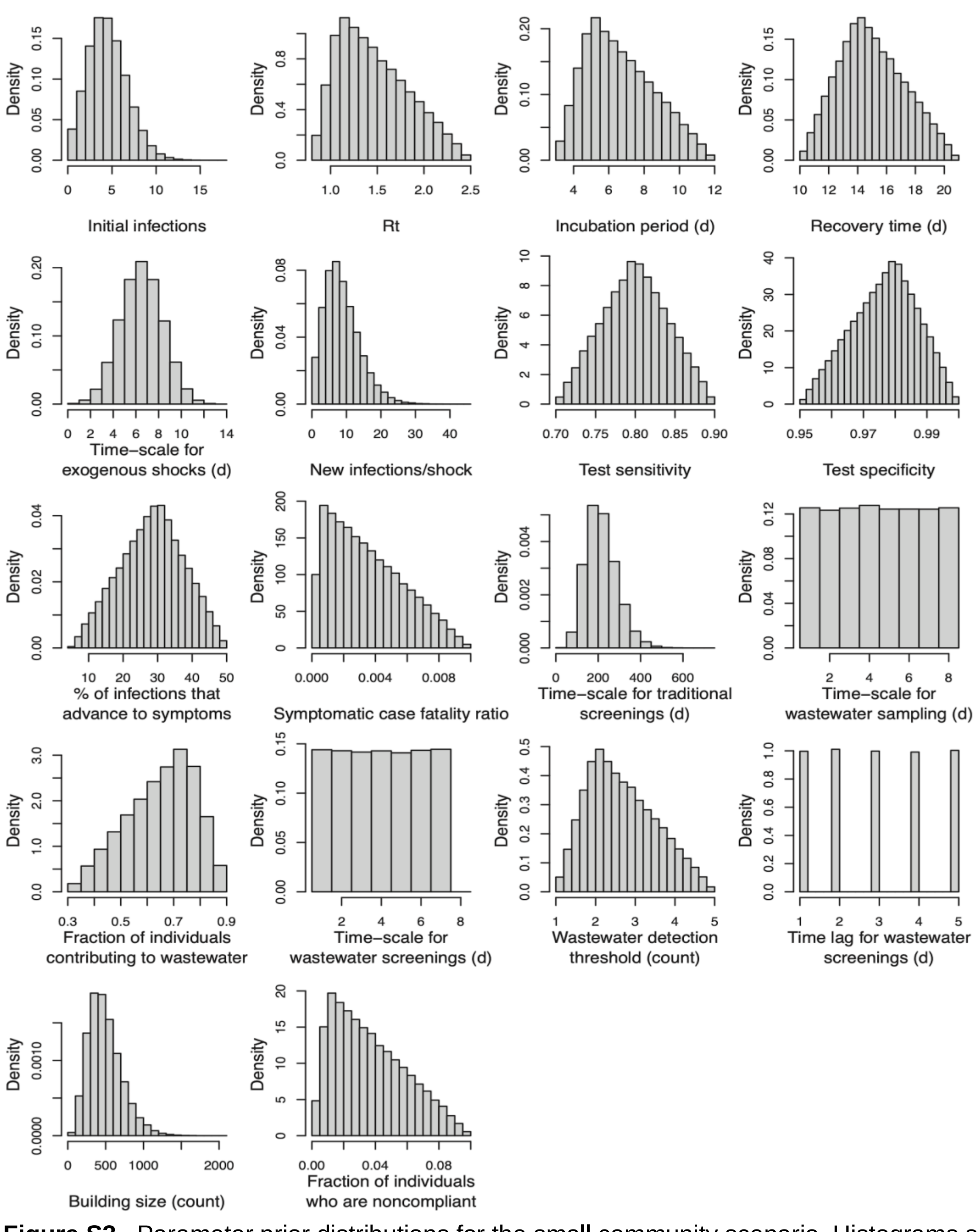


**Figure S3.** Parameter prior distributions for the small community scenario. Histograms show 100,000 samples from each parameter's marginal prior distribution.

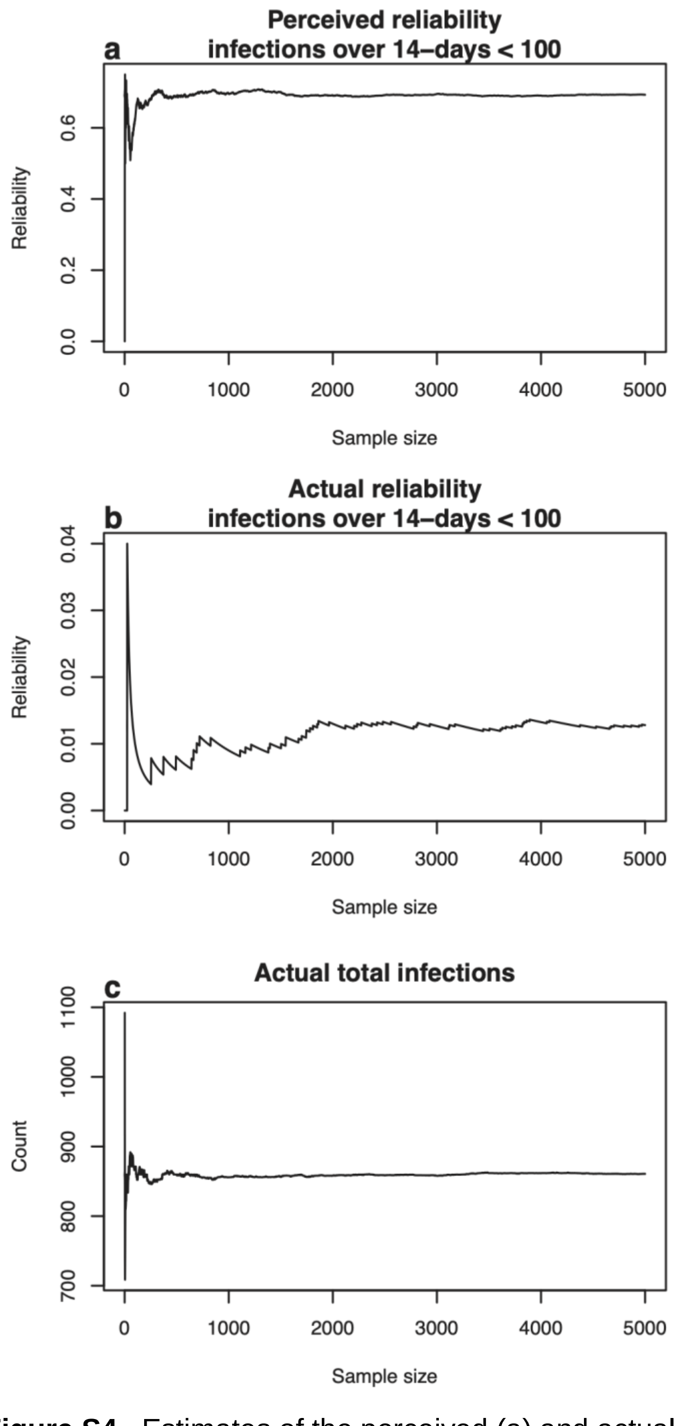


**Figure S4.** Estimates of the perceived (a) and actual (b) reliability of maintaining fewer than 100 infections across any 14-day period, and total number of infections (c) using subsamples of varying length. The results shown are using a 1-day lag before initiating wastewater-triggered screening tests and a 4-day turnaround time for completing the screening tests. The scenario includes $R_t$=1.1, 30 exogenous infections each week, and 10% noncompliance with quarantine procedures.

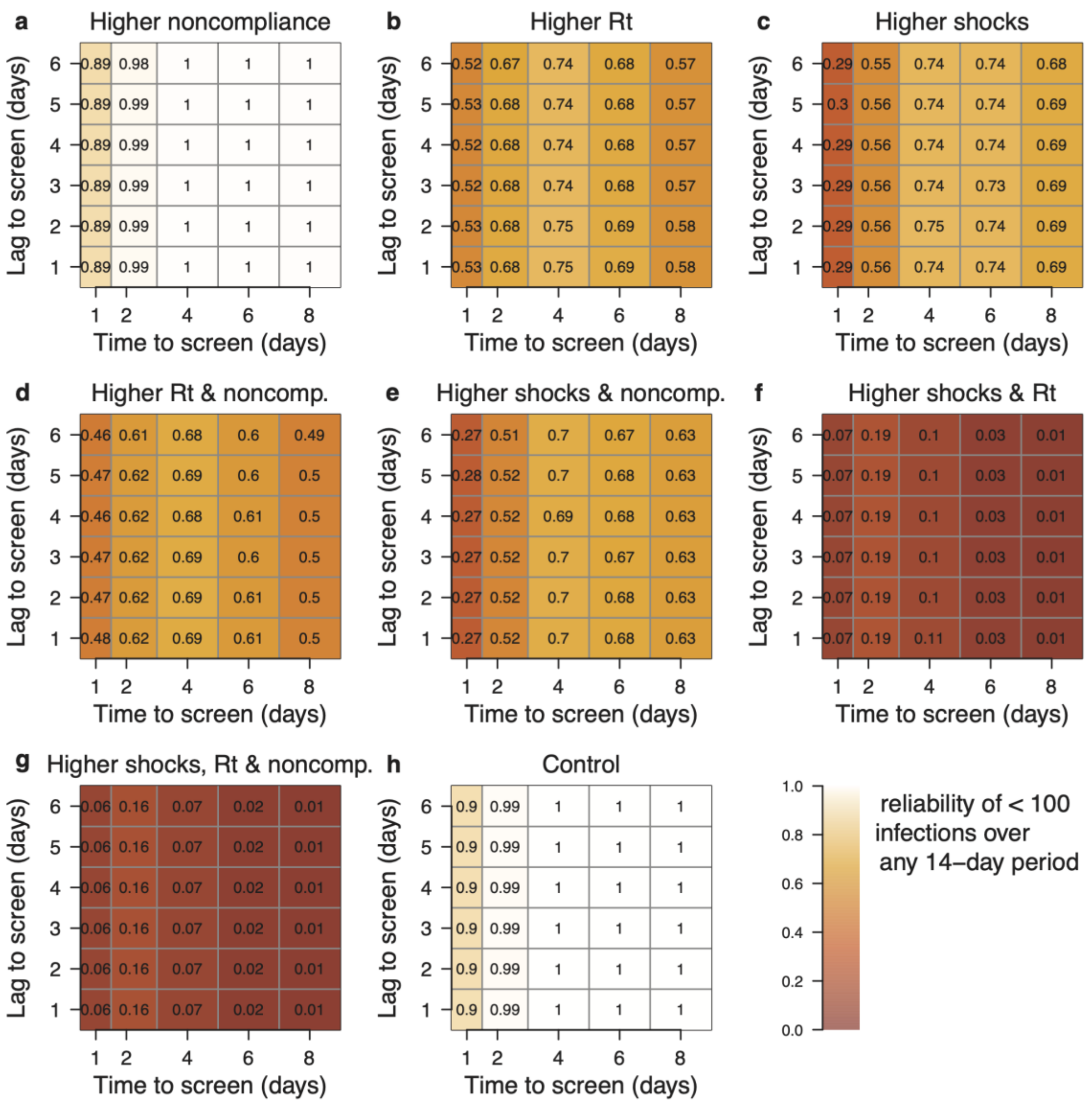

**Figure S5.** Reliability (probability) of maintaining fewer than 100 total new infections across any given 14-day period under different scenarios for wastewater-triggered screening tests. The reliability calculated here employs the perceived number of infections (false positive, true positive, and symptomatic cases) as opposed to the actual number of infections (asymptomatic, true positive and symptomatic cases), which corresponds to Fig 4 in the main text. A testing scenario includes the time required to conduct all screening tests (horizontal axis) and the lag time between when the wastewater sample results are received and when testing begins (vertical axis). The numbers given within each grid give the reliability, with lighter shading corresponding to higher reliability. All scenarios are named relative to the control scenario (h) in which $R_t$=1.1, there are weekly 15-exposure shocks, and 99% compliance with quarantine procedures. Shown are the cases where (a) the noncompliant fraction is increased to 10%; (b) $R_t$ is increased to 1.5; (c) weekly shocks are increased to 30 new exposures; (d) $R_t$=1.5 and $f_{nc}$ is increased to 10%; (e) shocks are worsened to 30 new exposures and $f_{nc}$=10%; (f) $R_t$=1.5 and shocks are worsened to 30 new exposures; (g) $R_t$=1.5, $f_{nc}$=10%, and weekly shocks are increased to 30 new exposures.

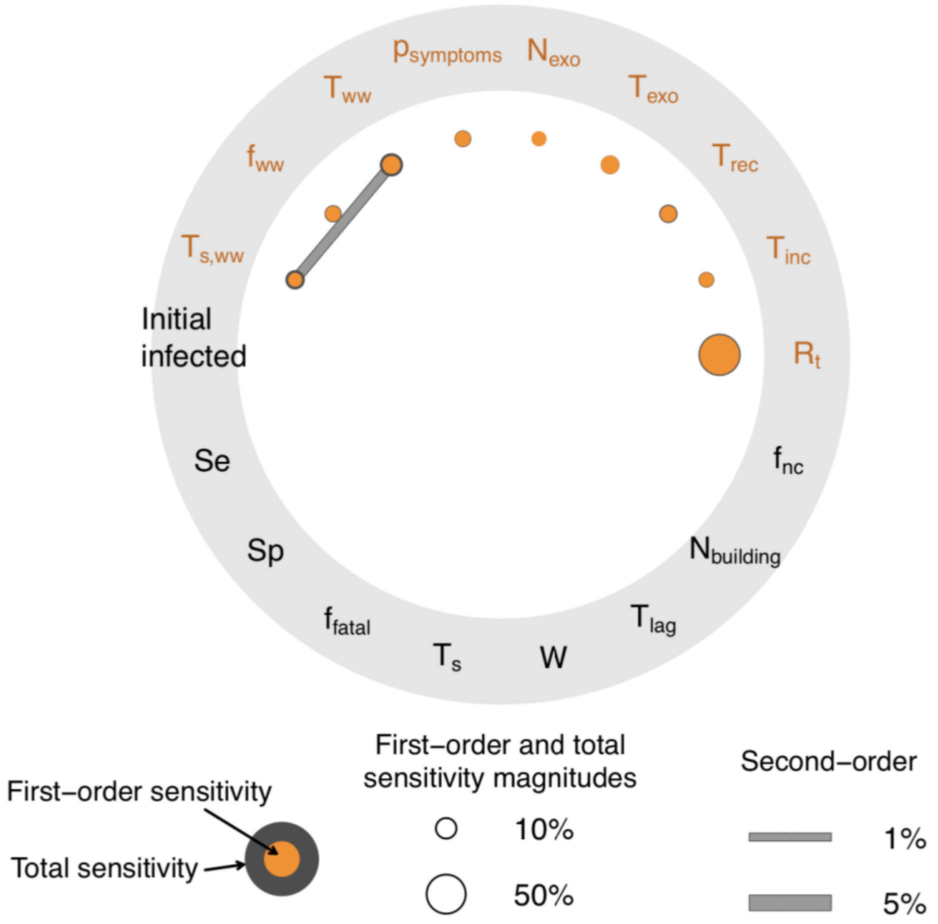


**Figure S6.** Sobol' radial convergence diagram depicting the decomposition of variance in the total number of infections among the uncertain input parameters for the high-risk scenario. Input parameter prior distributions have been chosen to represent the conditions and characteristics of a large university (20,000 campus community members) with higher rates of transmission (centered around $R_t$=2), more exogenous infections (about 14-50 new exposures twice per week), and higher rates of noncompliance (mode at 2%, but up to 15% noncompliance is possible with the prescribed prior distribution). Filled orange nodes represent first-order sensitivity indices (direct parameter influences); the concentric gray nodes represent total sensitivity indices (the given parameter's influence in combination with all other parameters); and filled gray bars represent second-order sensitivity indices for the interaction between the given pair of parameters. Parameters whose names or symbols are orange are the parameters to which the model is sensitive (in the upper hemisphere of the diagram); parameters in black (lower hemisphere) are those to which the model does not display a significant sensitivity.

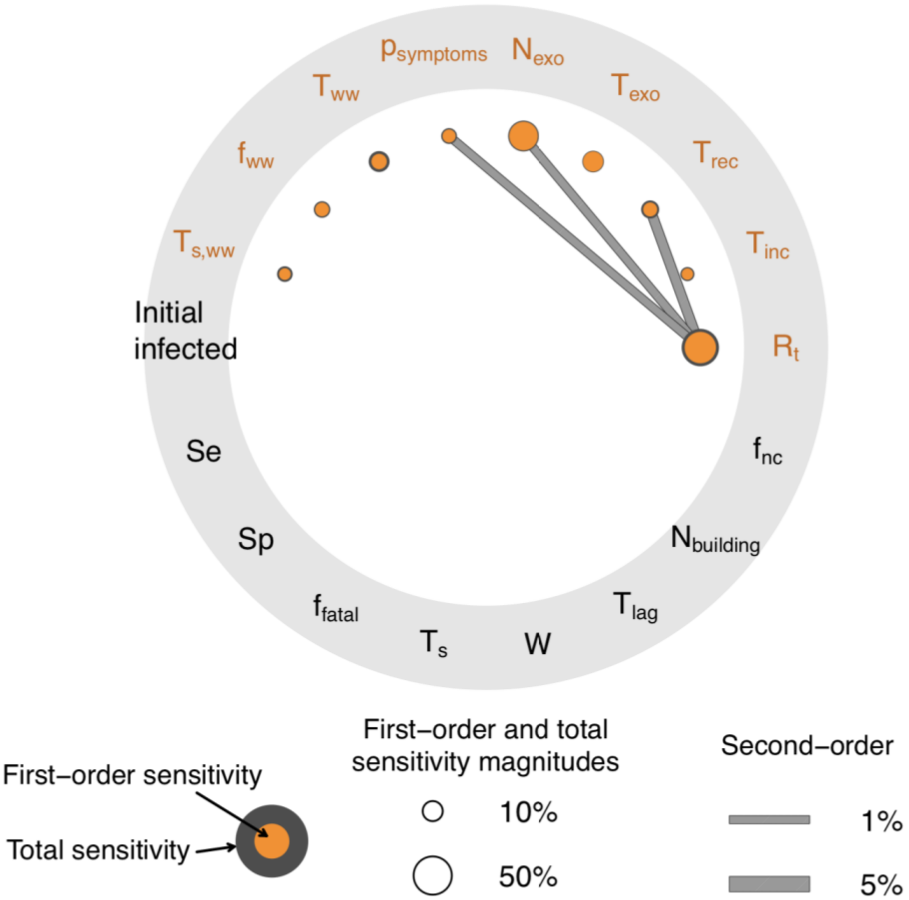


**Figure S7.** Sobol' radial convergence diagram depicting the decomposition of variance in the total number of infections among the uncertain input parameters for the small-college scenario. Input parameter prior distributions have been chosen to represent the conditions and characteristics of a small college (3,000 campus community members) with relatively low rates of transmission (centered around $R_t$=1.1), few exogenous infections (about 2-20 new exposures each week), and lower rates of noncompliance (mode at 1%, but up to 10% noncompliance is possible with the prescribed prior distribution). Filled orange nodes represent first-order sensitivity indices (direct parameter influences); the concentric gray nodes represent total sensitivity indices (the given parameter's influence in combination with all other parameters); and filled gray bars represent second-order sensitivity indices for the interaction between the given pair of parameters. Parameters whose names or symbols are orange are the parameters to which the model is sensitive (in the upper hemisphere of the diagram); parameters in black (lower hemisphere) are those to which the model does not display a significant sensitivity.

| Lag to screen (days) | 1 | 2 | 4 | 6 | 8 |
|---|---|---|---|---|---|
| 6 | 399 | 404 | 426 | 465 | 508 |
| 5 | 398 | 403 | 424 | 464 | 506 |
| 4 | 396 | 402 | 423 | 462 | 506 |
| 3 | 395 | 401 | 422 | 461 | 504 |
| 2 | 394 | 399 | 421 | 460 | 503 |
| 1 | 394 | 399 | 420 | 459 | 502 |

Time to screen (days)

**Table S1.** Cumulative infections for the control risk analysis scenario with $R_t$=1.1, 15 exogenous infections each week, and 1% noncompliance with quarantine procedures. Corresponds to Fig S5h.

| Lag to screen (days) | 1 | 2 | 4 | 6 | 8 |
|---|---|---|---|---|---|
| 6 | 419 | 425 | 449 | 492 | 538 |
| 5 | 418 | 424 | 448 | 491 | 537 |
| 4 | 417 | 423 | 447 | 490 | 536 |
| 3 | 416 | 421 | 446 | 488 | 534 |
| 2 | 415 | 420 | 444 | 487 | 533 |
| 1 | 415 | 420 | 443 | 486 | 532 |

Time to screen (days)

**Table S2.** Cumulative infections for the control risk analysis scenario with $R_t$=1.1, 15 exogenous infections each week, and 10% noncompliance with quarantine procedures. Corresponds to Fig S5a.

| Lag to screen (days) | 1 | 2 | 4 | 6 | 8 |
|---|---|---|---|---|---|
| 6 | 589 | 604 | 659 | 771 | 891 |
| 5 | 586 | 600 | 656 | 767 | 887 |
| 4 | 583 | 596 | 653 | 763 | 883 |
| 3 | 581 | 594 | 650 | 760 | 880 |
| 2 | 579 | 591 | 647 | 758 | 877 |
| 1 | 578 | 590 | 645 | 755 | 875 |

Time to screen (days)

**Table S3.** Cumulative infections for the control risk analysis scenario with $R_t$=1.5, 15 exogenous infections each week, and 1% noncompliance with quarantine procedures. Corresponds to Fig S5b.

| Lag to screen (days) | | | | | |
|---|---|---|---|---|---|
| 6 | 774 | 785 | 823 | 897 | 977 |
| 5 | 773 | 782 | 822 | 895 | 974 |
| 4 | 771 | 781 | 819 | 893 | 973 |
| 3 | 769 | 779 | 817 | 892 | 972 |
| 2 | 768 | 778 | 816 | 891 | 970 |
| 1 | 768 | 777 | 815 | 889 | 969 |
| Time to screen (days) | 1 | 2 | 4 | 6 | 8 |

**Table S4.** Cumulative infections for the control risk analysis scenario with $R_t$=1.1, 30 exogenous infections each week, and 1% noncompliance with quarantine procedures. Corresponds to Fig S5c.

| Lag to screen (days) | | | | | |
|---|---|---|---|---|---|
| 6 | 642 | 657 | 724 | 848 | 983 |
| 5 | 638 | 652 | 720 | 844 | 978 |
| 4 | 635 | 649 | 717 | 841 | 975 |
| 3 | 632 | 646 | 713 | 838 | 970 |
| 2 | 630 | 643 | 710 | 835 | 968 |
| 1 | 630 | 642 | 707 | 831 | 964 |
| Time to screen (days) | 1 | 2 | 4 | 6 | 8 |

**Table S5.** Cumulative infections for the control risk analysis scenario with $R_t$=1.5, 15 exogenous infections each week, and 10% noncompliance with quarantine procedures. Corresponds to Fig S5d.

| Lag to screen (days) | | | | | |
|---|---|---|---|---|---|
| 6 | 812 | 822 | 867 | 947 | 1030 |
| 5 | 811 | 821 | 866 | 946 | 1029 |
| 4 | 809 | 819 | 864 | 944 | 1027 |
| 3 | 807 | 817 | 861 | 943 | 1026 |
| 2 | 807 | 816 | 860 | 941 | 1024 |
| 1 | 807 | 815 | 859 | 940 | 1023 |
| Time to screen (days) | 1 | 2 | 4 | 6 | 8 |

**Table S6.** Cumulative infections for the control risk analysis scenario with $R_t$=1.1, 30 exogenous infections each week, and 10% noncompliance with quarantine procedures. Corresponds to Fig S5e.

| Lag to screen (days) | | | | | |
|---|---|---|---|---|---|
| 6 | 1120 | 1144 | 1249 | 1444 | 1654 |
| 5 | 1117 | 1139 | 1244 | 1440 | 1648 |
| 4 | 1113 | 1136 | 1240 | 1435 | 1642 |
| 3 | 1109 | 1133 | 1235 | 1432 | 1639 |
| 2 | 1108 | 1131 | 1231 | 1428 | 1635 |
| 1 | 1108 | 1129 | 1227 | 1425 | 1632 |
| | 1 | 2 | 4 | 6 | 8 |

Time to screen (days)

**Table S7.** Cumulative infections for the control risk analysis scenario with $R_t$=1.5, 30 exogenous infections each week, and 1% noncompliance with quarantine procedures. Corresponds to Fig S5f.

| Lag to screen (days) | | | | | |
|---|---|---|---|---|---|
| 6 | 1214 | 1240 | 1357 | 1579 | 1811 |
| 5 | 1210 | 1235 | 1353 | 1574 | 1807 |
| 4 | 1207 | 1231 | 1349 | 1570 | 1802 |
| 3 | 1204 | 1227 | 1345 | 1566 | 1798 |
| 2 | 1202 | 1224 | 1343 | 1562 | 1794 |
| 1 | 1202 | 1224 | 1341 | 1559 | 1790 |
| | 1 | 2 | 4 | 6 | 8 |

Time to screen (days)

**Table S8.** Cumulative infections for the control risk analysis scenario with $R_t$=1.5, 30 exogenous infections each week, and 10% noncompliance with quarantine procedures. Corresponds to Fig S5g.
